# Social innovation research checklist: A crowdsourcing open call and digital hackathon to develop a checklist for research to advance social innovation in health

**DOI:** 10.1101/2020.11.03.20225110

**Authors:** Eneyi Kpokiri, Elizabeth Chen, Jingjing Li, Sarah Payne, Priyanka Shrestha, Kaosar Afsana, Uche Amazigo, Phyllis Awor, Jean-Francois de Lavison, Saqif Khan, Jana D. Mier-Alpaño, Alberto Ong, Shivani Subhedar, Isabelle Wachmuth, Kala M. Mehta, Beatrice Halpaap, Joseph D. Tucker

**Author notes:** Co-first authors (Eneyi Kpokiri, Elizabeth Chen). Co-senior authors (Kala M. Mehta, Beatrice Halpaap, Joseph D. Tucker). Correspondence to: Joseph D. Tucker, 130 Mason Farm Road, Bioinformatics Building, University of North Carolina at Chapel Hill, Chapel Hill, NC, USA.

## Abstract

While social innovations in health have shown promise in closing the healthcare delivery gap, especially in low- and middle-income countries (LMICs), more research is needed to evaluate, scale up, and sustain social innovations. Research checklists can standardize and improve reporting of research findings, promote transparency, and increase replicability of study results and findings. This article describes the development of a 17-item social innovation in health research checklist to assess and report social innovation projects and provides examples of good reporting. The checklist is adapted from the TIDieR checklist and will facilitate more complete and transparent reporting and increase end user engagement.

**Summary points:**

- While many social innovations have been developed and shown promise in closing the healthcare delivery gap, more research is needed to evaluate social innovation
- The Social Innovation in Health Research Checklist, the first of its kind, is a 17-item checklist to improve reporting completeness and promote transparency in the development, implementation, and evaluation of social innovations in health
- The research checklist was developed through a three-step process, including a global open call for ideas, a scoping review, and a three-round modified Delphi process
- Use of this research checklist will enable researchers, innovators and partners to learn more about the process and results of social innovation in health research

## Introduction

Social innovations in health are inclusive solutions to address the healthcare delivery gap that meet the needs of end users through a multi-stakeholder, community-engaged process.(1) Many social innovations have been developed in response to specific community-based health needs. A subset has transformed healthcare in remote settings within low- and middle-income countries (LMICs). For example, social innovations have expanded private sector pharmacy-based services to manage childhood illnesses in Uganda(1), designed eco-health and community-based approach for Chagas control in Guatemala,(2) and increased gonorrhoea and chlamydia testing among sexual minorities in China.(3) While these social innovations have shown promise, research is needed to test, implement, adapt and scale up innovations and their impact.(1)

Research checklists provide one way to formalize and standardize reporting of research findings. Research checklists can spur multi-disciplinary research,(4,5) increase transparency,(4,6,7) improve reporting completeness(4,6,8) and facilitate easier comparison and replicability of study results and findings.(4,8,9) While some checklists are focused on reporting methods(9) and others focus more on the details in reporting results,(8) there are some checklists that report on both methods and results.(6) Overall, these checklists help researchers plan, execute, and report their processes and outcomes. However, there has been no research checklist targeting research for social innovation and only one focuses on design in global health.(4) In addition, meetings led by the Social Innovation in Health Initiative (SIHI), a group convened by TDR (the Special Programme for Research and Training in Tropical Diseases, co-sponsored by UNICEF, UNDP, the World Bank, and WHO), highlight the need for research tools to advance social innovation in healthcare delivery in LMICs.(10–12) The purpose of this manuscript is to describe the development of a research checklist to assess and report social innovation projects as well as highlight the importance of research in social innovation projects.

## Methods

Our working group used a three-step process, including an open call for ideas, a scoping review, and a modified Delphi process. This three-step process resulted in the development of a Social Innovation in Health Research Checklist as well as a Social Innovation in Health Monitoring and Evaluation Framework.(13)

### Open call

Social Entrepreneurship to Spur Health (SESH) is the research hub in China within the TDR SIHI. SESH and SIHI jointly organized a global crowdsourcing open call to solicit creative ideas and tools on the development of a social innovation research checklist, as well as ideas on measuring social innovation in health performance to develop a conceptual framework for measurement and evaluation. The purpose of the checklist was to develop a list of key components related to social innovation in health research. The measurement ideas were to help project managers and their teams effectively implement their social innovation projects, guide and improve project design and allow them to more accurately report and measure the impact of their projects.

We formed a steering committee to finalize the call for submissions, decide the prize structure, identify judges, and advise on implementation. Steering committee members for this open call included researchers, innovators, policy makers, implementers, and students. This process was similar to other crowdsourcing open calls organized by SESH to understand research mentorship in LMICs(14) and to promote HIV testing and hepatitis testing where online open calls led to in-person consensus-building meetings for further action.(15,16)

The open call was launched in November 2019 and closed in February 2020. During this time, the open call was distributed within the SIHI network, through social media channels (e.g. Twitter), on SESH’s website, and through other partner and academic networks. The open call solicited monitoring and evaluation frameworks, research checklists, and methods for assessing monitoring and evaluation. Eligibility criteria included written in English, less than 1,000 words, and focused on monitoring and evaluation. Volunteer judges were selected, with a focus on people in LMICs who have experience in social innovation. After the open call was closed, each submission was screened independently for eligibility and eligible entries were reviewed by five independent judges.

### Scoring entries

Entries were judged in three categories: (1) relevance to inform a standardized framework or research checklist, (2) creativity, and (3) the participant’s experience in the field of social innovation. Scores were assigned between “1” and “10” in each category and then averaged for a final score of the entry. Entries that achieved a mean score of “7” and above were deemed semi-finalists. Semi-finalists entries were then reviewed once more by the steering committee, and finalists were selected. Finalist submissions were chosen by the steering committee in March 2020 and invited to join a hackathon to finalize the research checklist. Hackathons are a form of crowdsourcing that include an open call for participants, a sprint collaborative event, and follow-up activities.(17)

Given the COVID-19 pandemic, we transitioned our originally planned in-person workshop to a digital consensus-building process composed of three two-hour videoconferences. Instead of meeting in-person over three consecutive days, we scheduled videoconference workshops over the span of several weeks plus an additional videoconference focused on introductions and logistics. Further details about the hackathon’s digital consensus-building process are described in the section on the modified Delphi process below.

### Scoping review

The steering committee reviewed peer-reviewed literature and grey literature related to social innovation in health to understand the current landscape and existing research and practice efforts in this field.

### Modified Delphi process

The Delphi process is a structured method to develop consensus and is commonly used to develop health guidelines and research checklists.(18) A typical Delphi process has a group of experts iteratively develop a consensus. Given the importance of end users in social innovation, our Delphi process was modified to incorporate feedback from expert (three rounds) and end-users (two rounds). The expert group consisted of the steering committee and finalists from the crowdsourcing open call. The user group included people with experience and/or interest in social innovation research. Iterative feedback from each of the three Delphi surveys was used to revise the research checklist and monitoring and evaluation conceptual framework. Initial feedback focused on open responses to draft items and later rounds included close-ended Likert scale responses ranging from strongly agree to strongly disagree assessing whether we should include the different components of the research checklist.

## Results

### Open call

We received a total of 21 unique submissions from 12 different countries: United States of America (n=5), Bangladesh (n=3), Colombia (n=2), Nigeria (n=2), Philippines (n=2), Cameroon (n=1), Guinea (n=1), Honduras (n=1), India (n=1), Kenya (n=1), Thailand (n=1), and United Kingdom (n=1). Therefore 65% (11 out of 17) of the unique submissions (all those except entries submitted from the United States and the United Kingdom) were from LMICs. After the initial screening, 17 out of the 21 submissions were deemed eligible for judging. After the steering committee discussion, four finalists were selected: two from the United States, one from the Philippines, and one from Bangladesh.

We noted several themes across finalist entries, including the following: a strong focus on community and stakeholder engagement; considering implementation as an essential component; and examining financial models and financial sustainability.

### Modified Delphi process

The four workshops related to consensus development focused on the consensus-building process, ideas from open call finalists, the results of the scoping review, and preliminary content for the monitoring and evaluation conceptual framework and research checklist. In between videoconference meetings, steering committee members, finalists, and invited stakeholders were asked to complete online Qualtrics surveys as part of the modified Delphi process.

### Discussions at videoconference workshops

During each of our videoconference workshops, participants discussed potential components of the research checklist. For example, one of the major topics of discussion at our second meeting focused on the topic of financing and how sustainability and revenue generation activities are not consistently reported. The discussion uncovered that some participants felt that financing and sustainability should be explicitly included in the research checklist. We included this item in the draft research checklist and used the modified Delphi process to determine the content of the final version of the checklist.

### Delphi surveys

The first Delphi survey was completed by 65 out of 96 invited participants. Overall responses included structuring the preamble with mission statement and adding important definitions, specifying and clarifying each checklist item, defining terms used such as health, stakeholders, facilitators vs. providers, and open access resources. Feedback during the first few consensus-building videoconference meetings was further incorporated such as including additional items, limitations and strengths.

The second Delphi survey was conducted four weeks after the initial survey. It was completed by 22 out of 45 invited participants. An end-user meeting was also convened to solicit innovators perspective into the research checklist elements as a separate digital meeting. Further enhancement on each item of the checklist was done: descriptions of social innovation was added, consistency on using terminologies was ensured (end users vs beneficiaries), and descriptions of each were clarified.

The final survey by 16 out of 25 invited participants. Minor adjustments at this stage included fixing grammatical errors and harmonizing definitions.

### Social innovation in health research checklist

Our social innovation in health research checklist uses a variety of terms that are defined differently across disciplines. The social innovation research checklist is adapted from the TIDieR checklist that focuses on better reporting of interventions.(8) Key terms are defined in Table 1.

**Table 1.**
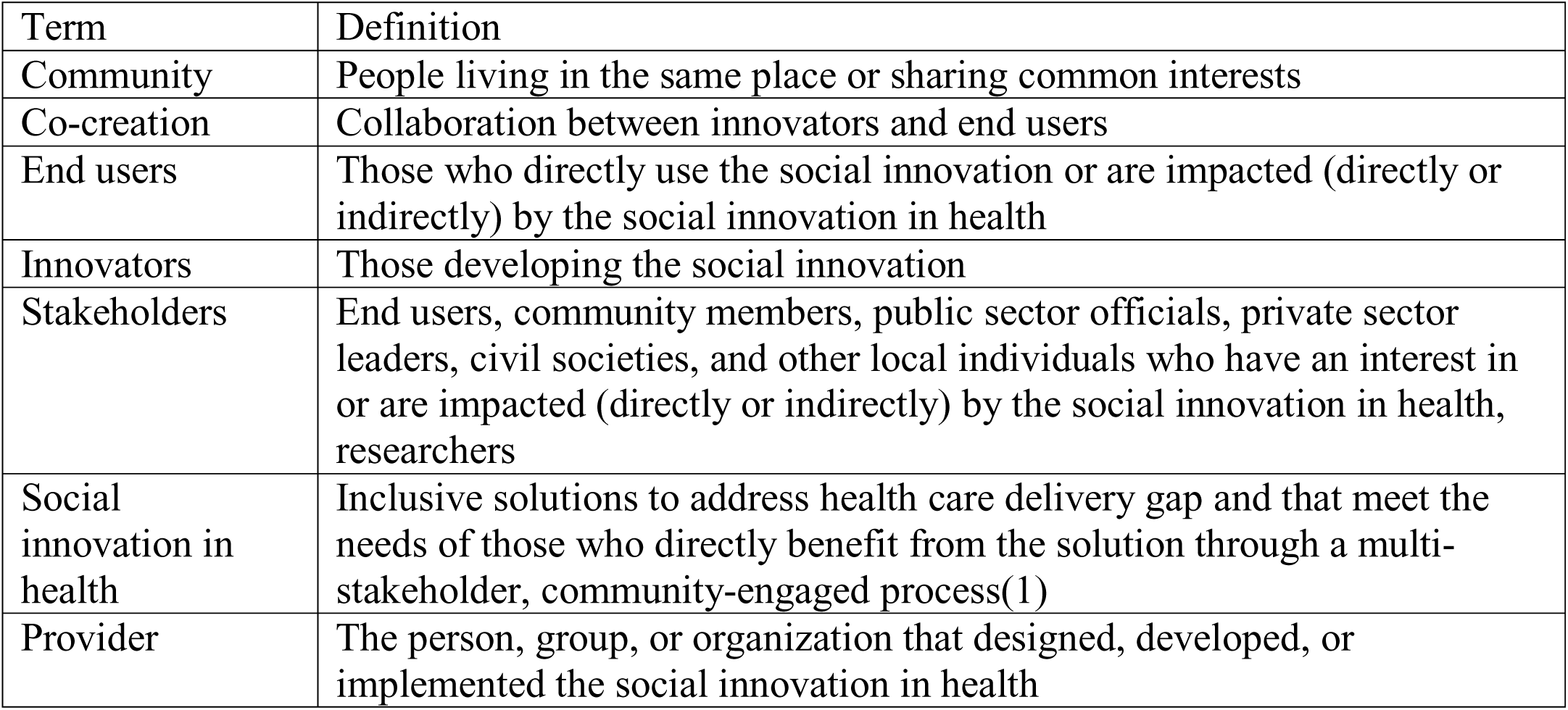
Terms and definitions for our social innovation in health research checklist

| Term | Definition |
| --- | --- |
| Community | People living in the same place or sharing common interests |
| Co-creation | Collaboration between innovators and end users |
| End users | Those who directly use the social innovation or are impacted (directly or indirectly) by the social innovation in health |
| Innovators | Those developing the social innovation |
| Stakeholders | End users, community members, public sector officials, private sector leaders, civil societies, and other local individuals who have an interest in or are impacted (directly or indirectly) by the social innovation in health, researchers |
| Social innovation in health | Inclusive solutions to address health care delivery gap and that meet the needs of those who directly benefit from the solution through a multi-stakeholder, community-engaged process(1) |
| Provider | The person, group, or organization that designed, developed, or implemented the social innovation in health |

At the end of our multi-step process, we finalized a research checklist with 17 items (Table 2). Table 2 includes the social innovation in health research checklist, a description of each of the items, and the percentage of Delphi survey respondents who affirmed that each item should be included in our final survey. We have also included a supplemental file with the checklist in PDF format along with a list of useful resources and additional information about the Social Innovation in Health Initiative research hubs. We gathered this set of resources from steering committee members and finalists during our checklist development process. In addition, we list three examples of a completed checklist in Table 3. They describe a social innovation research on Chagas disease in Guatemala,(2) maternal health in Uganda,(16) and sexual health in China.(3)

**Table 2.**
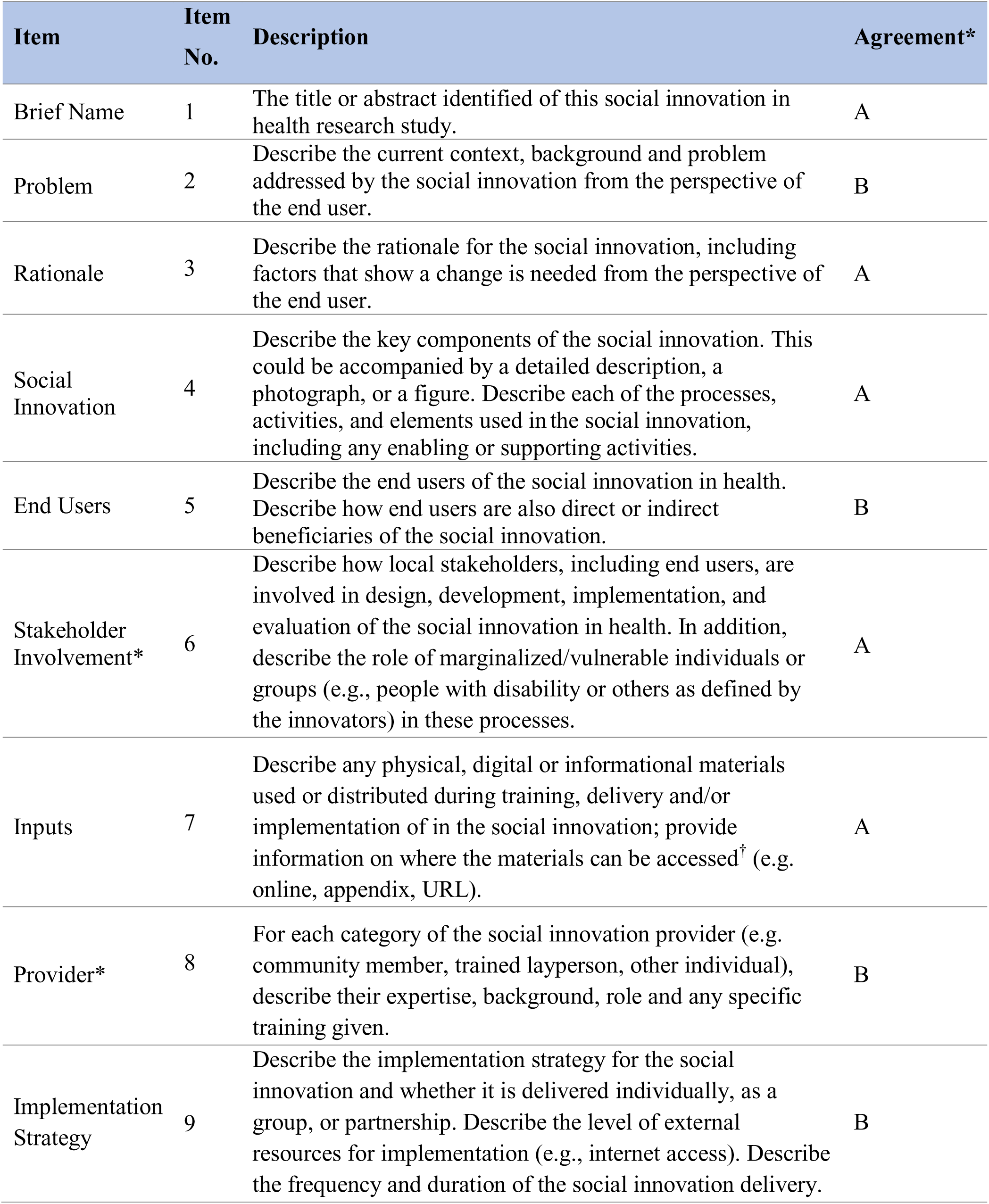
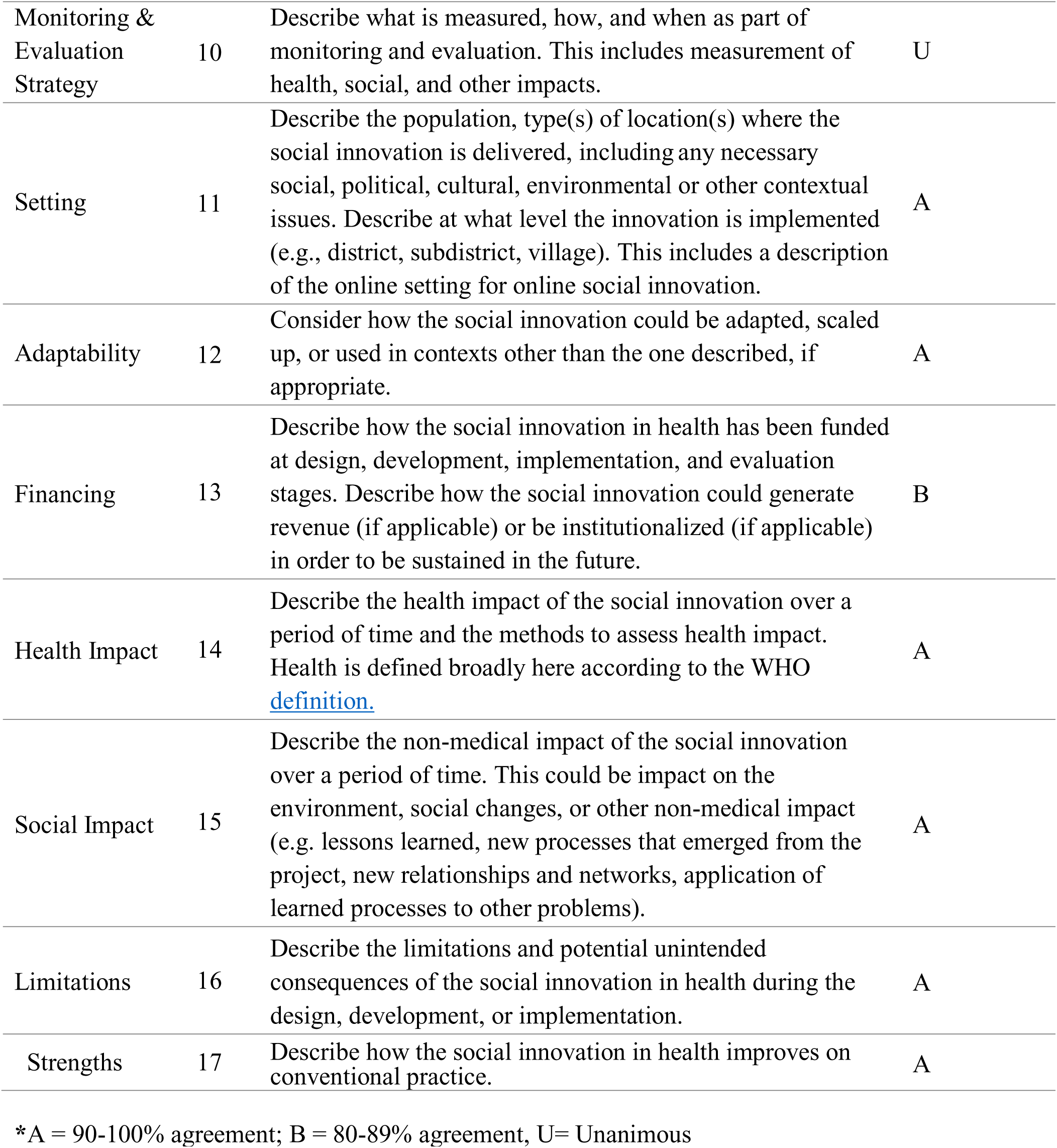
Social Innovation in Health Research Checklist

**Table 3.**
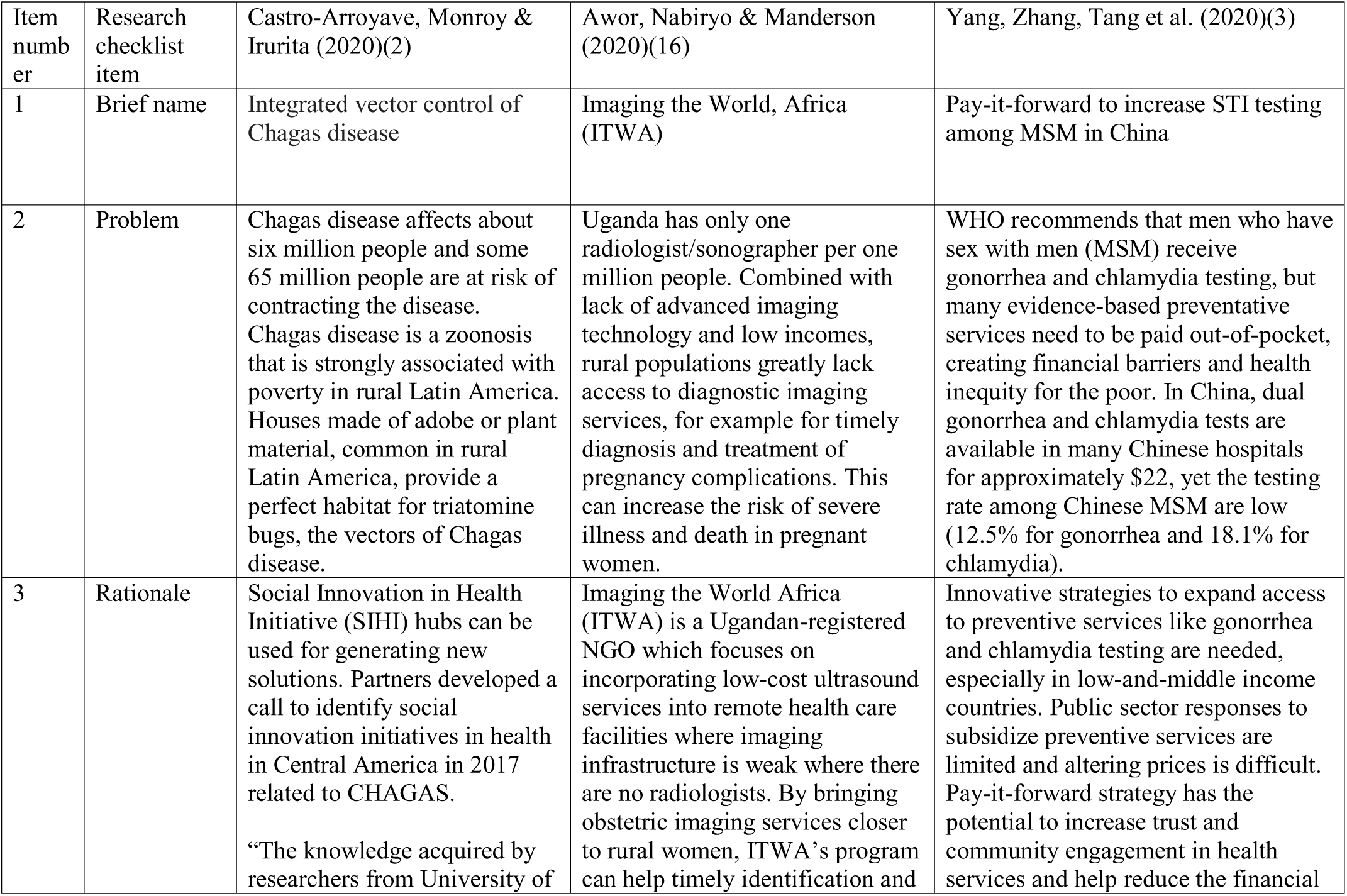
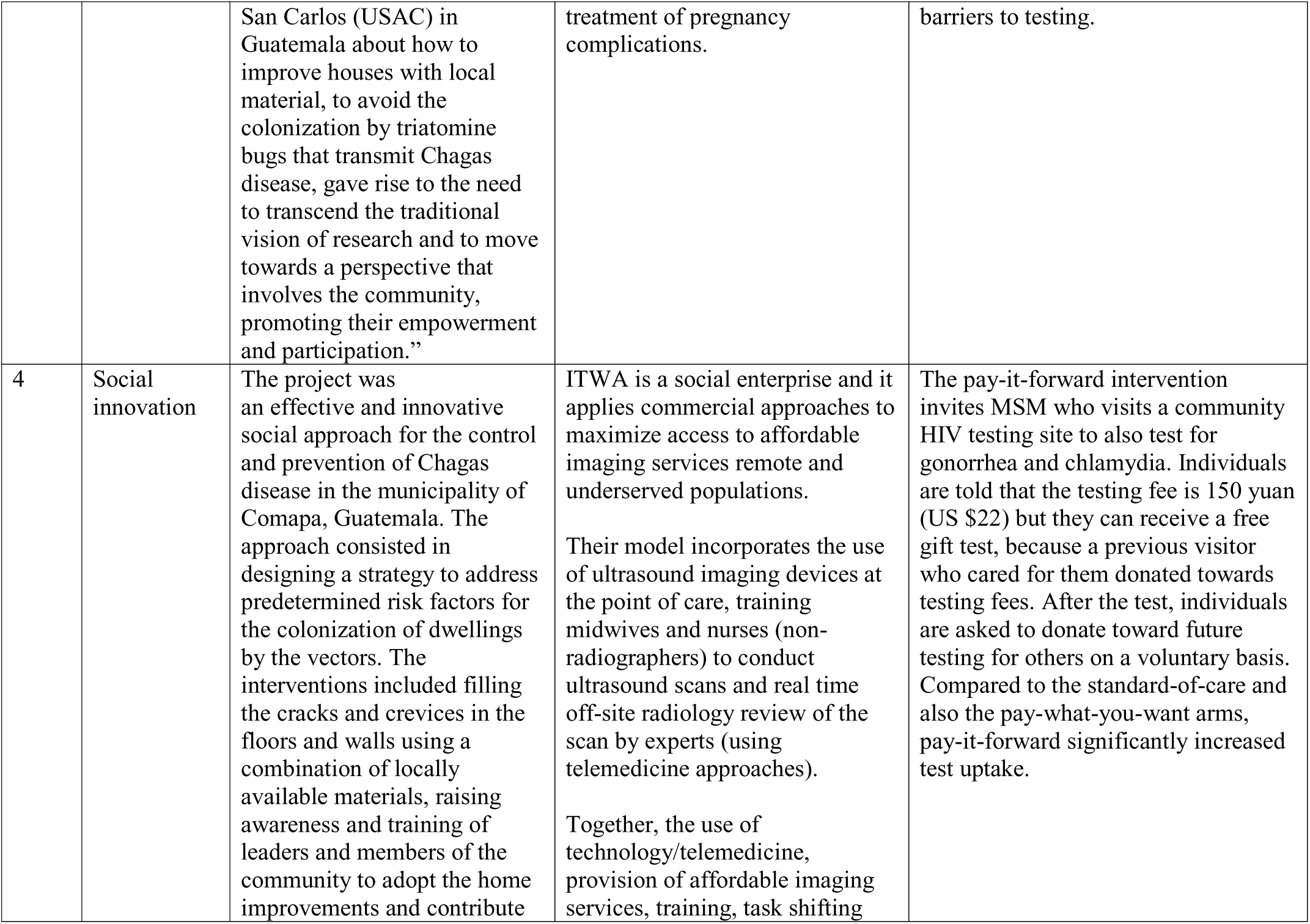
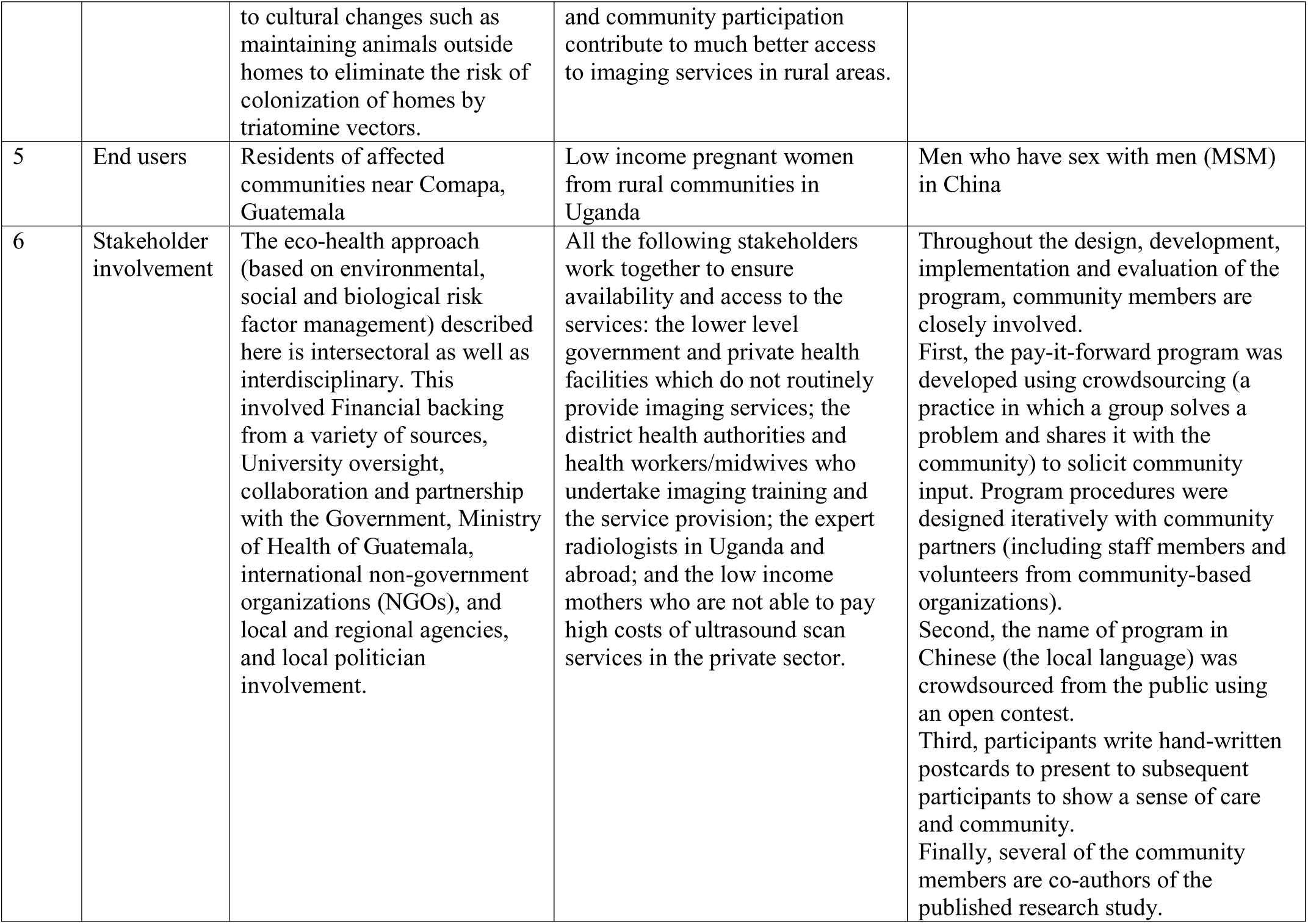
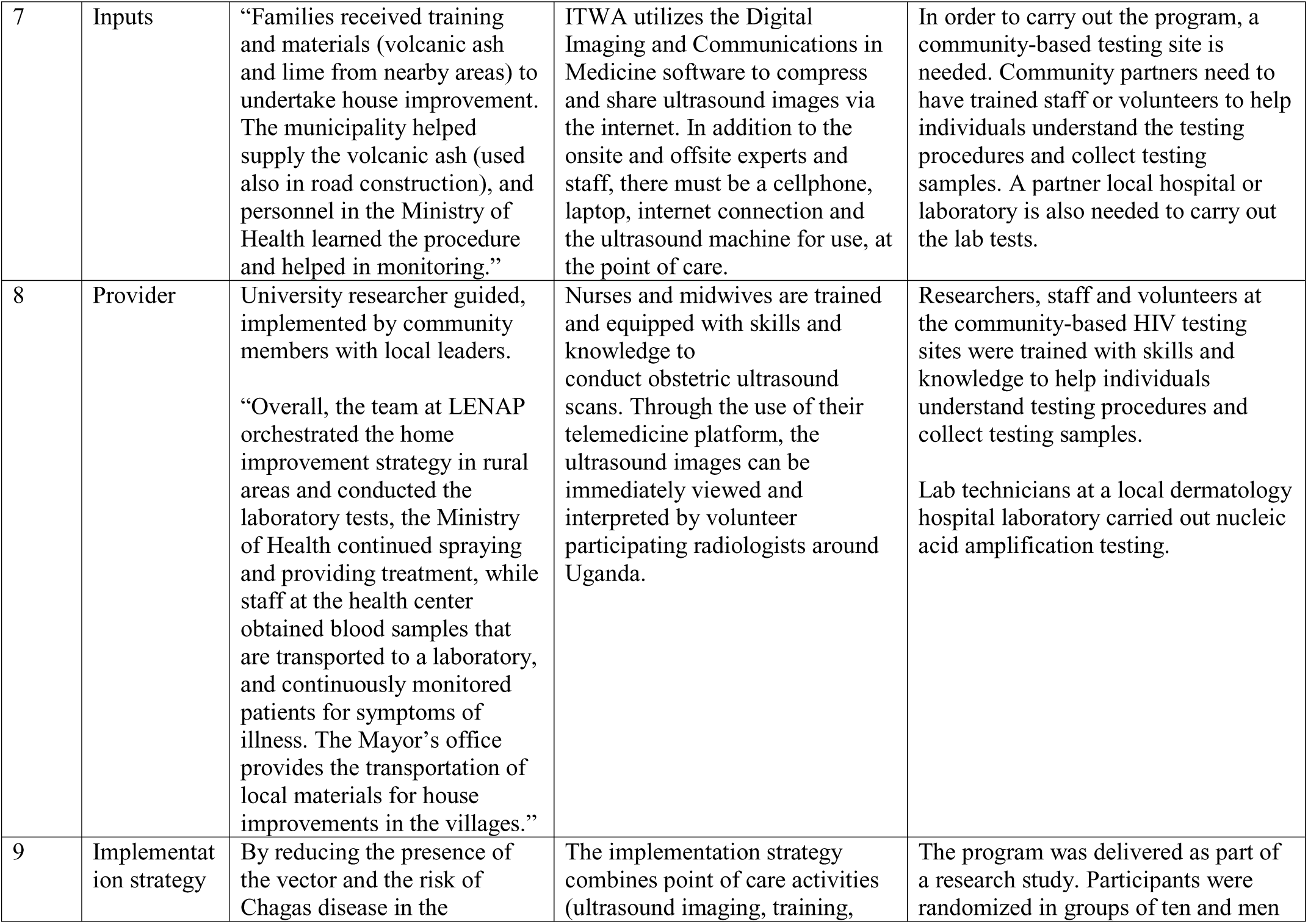
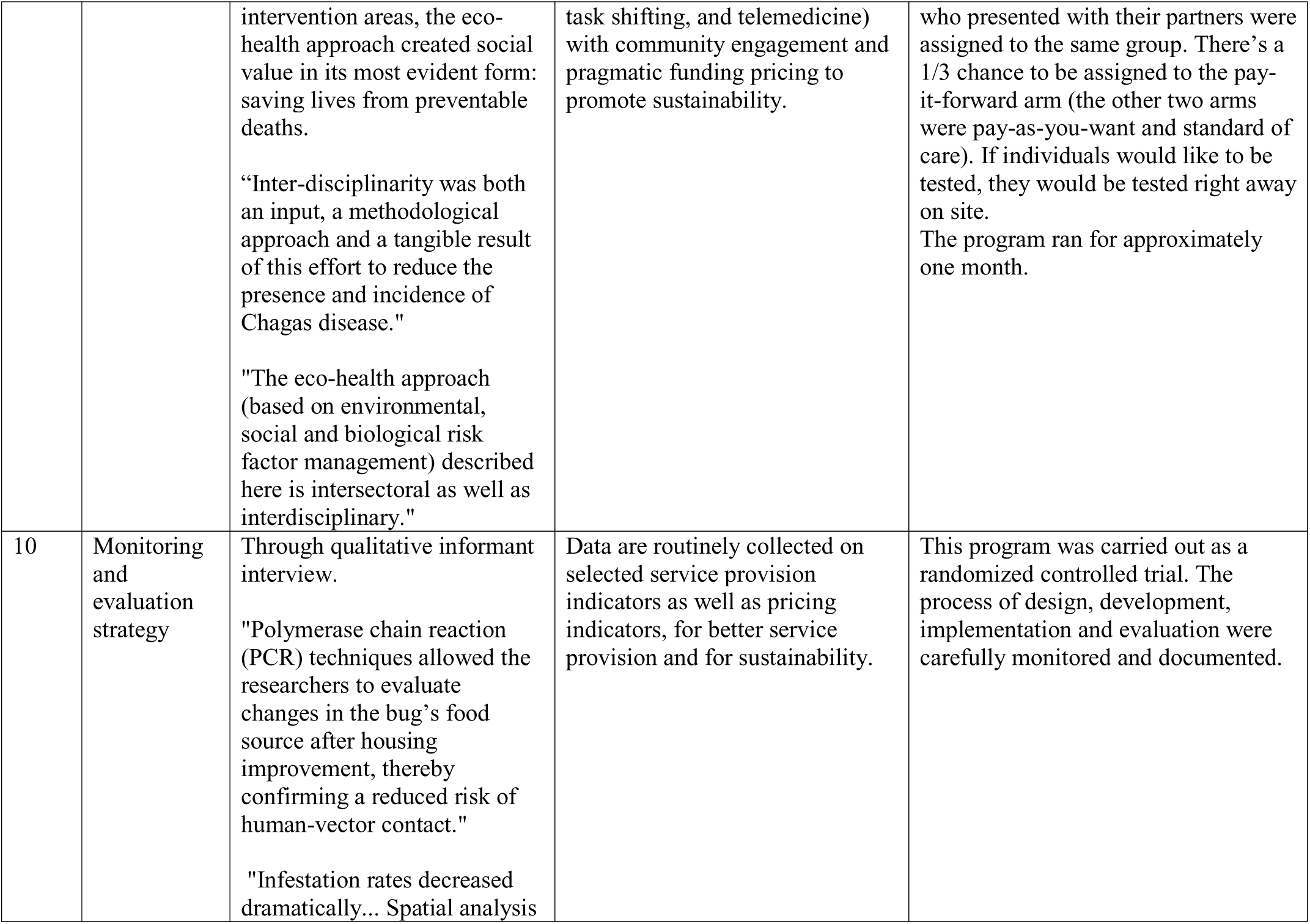
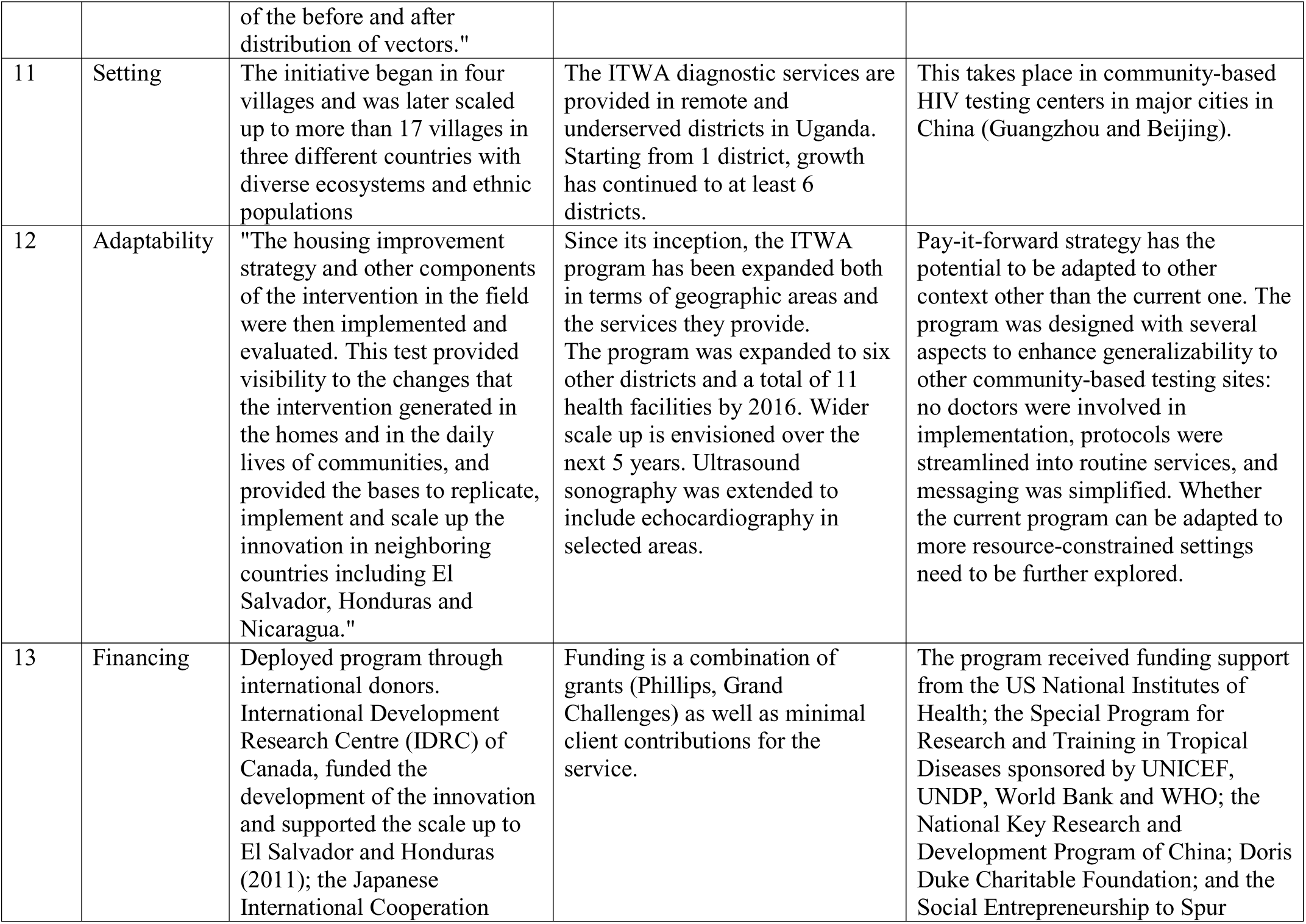
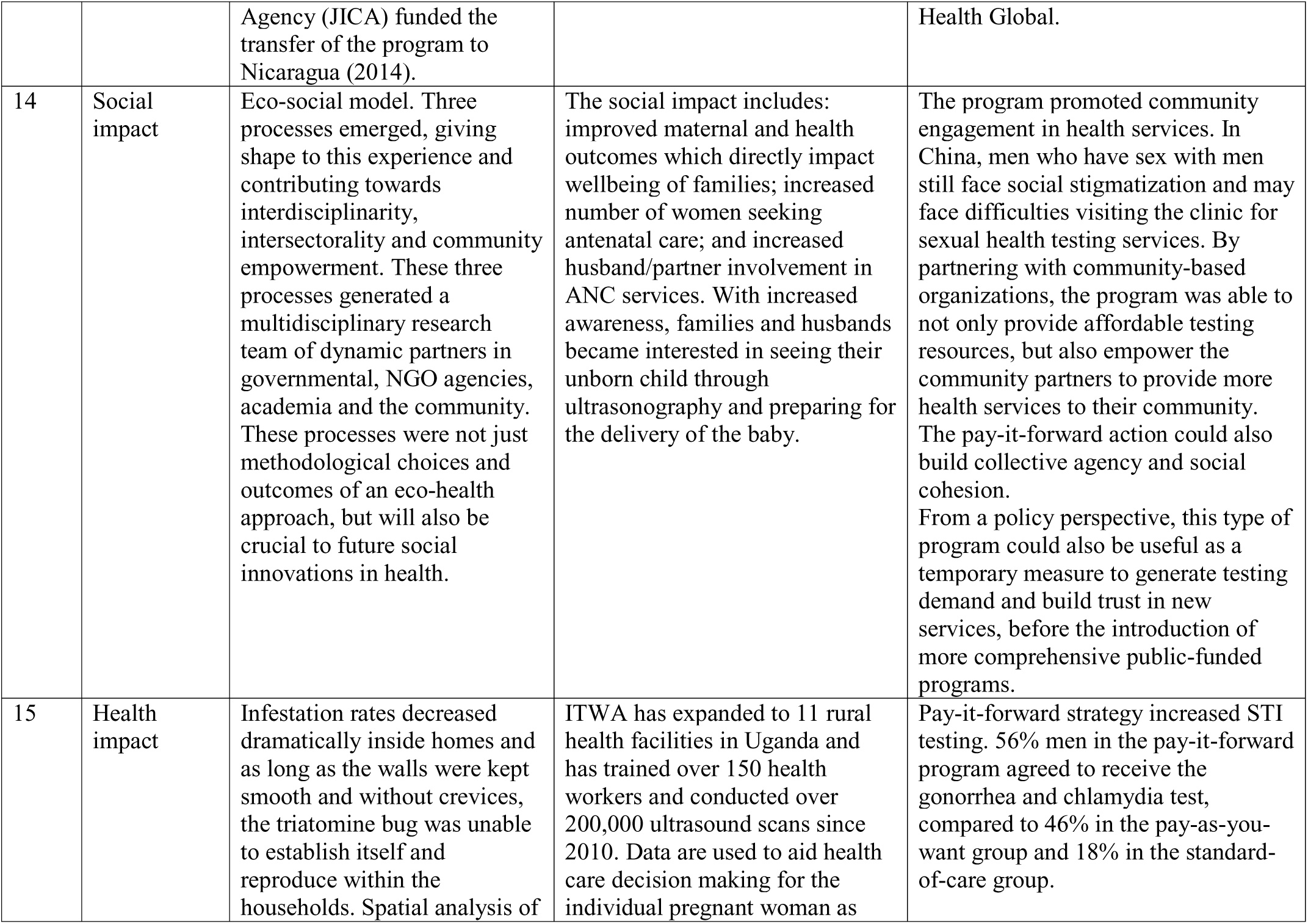
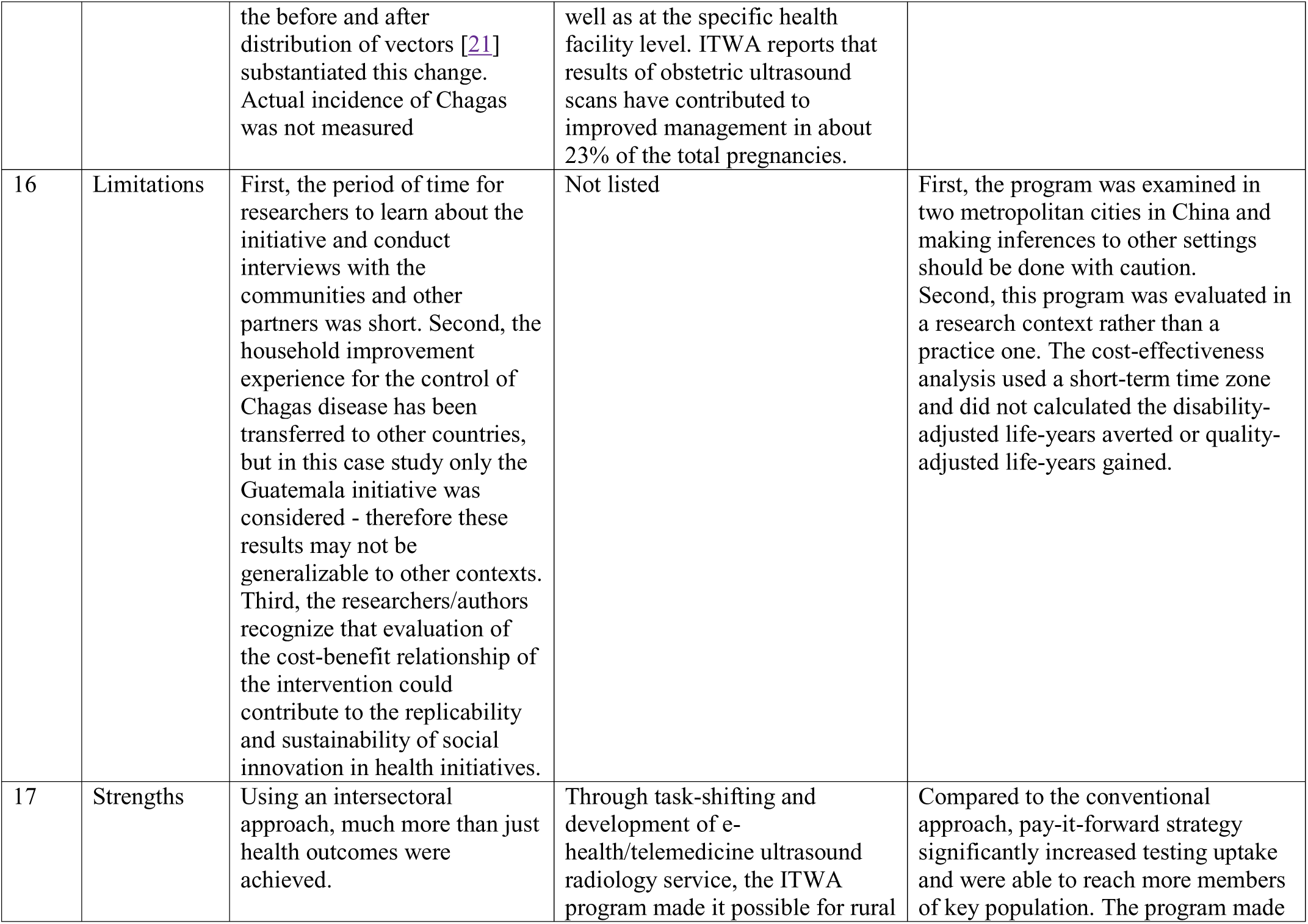
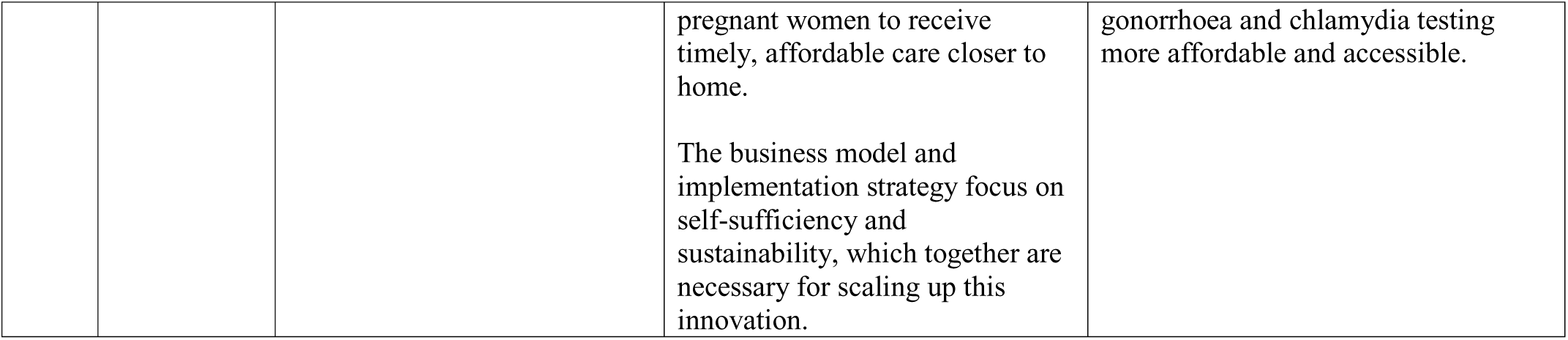
Examples of Social innovations in health described using the new research checklist

| Item number | Research checklist item | Castro-Arroyave, Monroy & Irurita (2020)(2) | Awor, Nabiryo & Manderson (2020)(16) | Yang, Zhang, Tang et al. (2020)(3) |
| --- | --- | --- | --- | --- |
| 1 | Brief name | Integrated vector control of Chagas disease | Imaging the World, Africa (ITWA) | Pay-it-forward to increase STI testing among MSM in China |
| 2 | Problem | Chagas disease affects about six million people and some 65 million people are at risk of contracting the disease. Chagas disease is a zoonosis that is strongly associated with poverty in rural Latin America. Houses made of adobe or plant material, common in rural Latin America, provide a perfect habitat for triatomine bugs, the vectors of Chagas disease. | Uganda has only one radiologist/sonographer per one million people. Combined with lack of advanced imaging technology and low incomes, rural populations greatly lack access to diagnostic imaging services, for example for timely diagnosis and treatment of pregnancy complications. This can increase the risk of severe illness and death in pregnant women. | WHO recommends that men who have sex with men (MSM) receive gonorrhea and chlamydia testing, but many evidence-based preventative services need to be paid out-of-pocket, creating financial barriers and health inequity for the poor. In China, dual gonorrhea and chlamydia tests are available in many Chinese hospitals for approximately \$22, yet the testing rate among Chinese MSM are low (12.5% for gonorrhea and 18.1% for chlamydia). |
| 3 | Rationale | Social Innovation in Health Initiative (SIHI) hubs can be used for generating new solutions. Partners developed a call to identify social innovation initiatives in health in Central America in 2017 related to CHAGAS.<br>“The knowledge acquired by researchers from University of | Imaging the World Africa (ITWA) is a Ugandan-registered NGO which focuses on incorporating low-cost ultrasound services into remote health care facilities where imaging infrastructure is weak where there are no radiologists. By bringing obstetric imaging services closer to rural women, ITWA’s program can help timely identification and | Innovative strategies to expand access to preventive services like gonorrhea and chlamydia testing are needed, especially in low-and-middle income countries. Public sector responses to subsidize preventive services are limited and altering prices is difficult. Pay-it-forward strategy has the potential to increase trust and community engagement in health services and help reduce the financial |
|  |  | San Carlos (USAC) in Guatemala about how to improve houses with local material, to avoid the colonization by triatomine bugs that transmit Chagas disease, gave rise to the need to transcend the traditional vision of research and to move towards a perspective that involves the community, promoting their empowerment and participation.” | treatment of pregnancy complications. | barriers to testing. |
| 4 | Social innovation | The project was an effective and innovative social approach for the control and prevention of Chagas disease in the municipality of Comapa, Guatemala. The approach consisted in designing a strategy to address predetermined risk factors for the colonization of dwellings by the vectors. The interventions included filling the cracks and crevices in the floors and walls using a combination of locally available materials, raising awareness and training of leaders and members of the community to adopt the home improvements and contribute | ITWA is a social enterprise and it applies commercial approaches to maximize access to affordable imaging services remote and underserved populations.<br>Their model incorporates the use of ultrasound imaging devices at the point of care, training midwives and nurses (non-radiographers) to conduct ultrasound scans and real time off-site radiology review of the scan by experts (using telemedicine approaches).<br>Together, the use of technology/telemedicine, provision of affordable imaging services, training, task shifting | The pay-it-forward intervention invites MSM who visits a community HIV testing site to also test for gonorrhea and chlamydia. Individuals are told that the testing fee is 150 yuan (US \$22) but they can receive a free gift test, because a previous visitor who cared for them donated towards testing fees. After the test, individuals are asked to donate toward future testing for others on a voluntary basis. Compared to the standard-of-care and also the pay-what-you-want arms, pay-it-forward significantly increased test uptake. |
|  |  | to cultural changes such as maintaining animals outside homes to eliminate the risk of colonization of homes by triatomine vectors. | and community participation contribute to much better access to imaging services in rural areas. |  |
| 5 | End users | Residents of affected communities near Comapa, Guatemala | Low income pregnant women from rural communities in Uganda | Men who have sex with men (MSM) in China |
| 6 | Stakeholder involvement | The eco-health approach (based on environmental, social and biological risk factor management) described here is intersectoral as well as interdisciplinary. This involved Financial backing from a variety of sources, University oversight, collaboration and partnership with the Government, Ministry of Health of Guatemala, international non-government organizations (NGOs), and local and regional agencies, and local politician involvement. | All the following stakeholders work together to ensure availability and access to the services: the lower level government and private health facilities which do not routinely provide imaging services; the district health authorities and health workers/midwives who undertake imaging training and the service provision; the expert radiologists in Uganda and abroad; and the low income mothers who are not able to pay high costs of ultrasound scan services in the private sector. | Throughout the design, development, implementation and evaluation of the program, community members are closely involved.<br>First, the pay-it-forward program was developed using crowdsourcing (a practice in which a group solves a problem and shares it with the community) to solicit community input. Program procedures were designed iteratively with community partners (including staff members and volunteers from community-based organizations).<br>Second, the name of program in Chinese (the local language) was crowdsourced from the public using an open contest.<br>Third, participants write hand-written postcards to present to subsequent participants to show a sense of care and community.<br>Finally, several of the community members are co-authors of the published research study. |
| 7 | Inputs | “Families received training and materials (volcanic ash and lime from nearby areas) to undertake house improvement. The municipality helped supply the volcanic ash (used also in road construction), and personnel in the Ministry of Health learned the procedure and helped in monitoring.” | ITWA utilizes the Digital Imaging and Communications in Medicine software to compress and share ultrasound images via the internet. In addition to the onsite and offsite experts and staff, there must be a cellphone, laptop, internet connection and the ultrasound machine for use, at the point of care. | In order to carry out the program, a community-based testing site is needed. Community partners need to have trained staff or volunteers to help individuals understand the testing procedures and collect testing samples. A partner local hospital or laboratory is also needed to carry out the lab tests. |
| 8 | Provider | University researcher guided, implemented by community members with local leaders.<br>“Overall, the team at LENAP orchestrated the home improvement strategy in rural areas and conducted the laboratory tests, the Ministry of Health continued spraying and providing treatment, while staff at the health center obtained blood samples that are transported to a laboratory, and continuously monitored patients for symptoms of illness. The Mayor’s office provides the transportation of local materials for house improvements in the villages.” | Nurses and midwives are trained and equipped with skills and knowledge to conduct obstetric ultrasound scans. Through the use of their telemedicine platform, the ultrasound images can be immediately viewed and interpreted by volunteer participating radiologists around Uganda. | Researchers, staff and volunteers at the community-based HIV testing sites were trained with skills and knowledge to help individuals understand testing procedures and collect testing samples.<br>Lab technicians at a local dermatology hospital laboratory carried out nucleic acid amplification testing. |
| 9 | Implementation strategy | By reducing the presence of the vector and the risk of Chagas disease in the | The implementation strategy combines point of care activities (ultrasound imaging, training, | The program was delivered as part of a research study. Participants were randomized in groups of ten and men |
|  |  | intervention areas, the eco-health approach created social value in its most evident form: saving lives from preventable deaths.<br>"Inter-disciplinarity was both an input, a methodological approach and a tangible result of this effort to reduce the presence and incidence of Chagas disease."<br>"The eco-health approach (based on environmental, social and biological risk factor management) described here is intersectoral as well as interdisciplinary." | task shifting, and telemedicine) with community engagement and pragmatic funding pricing to promote sustainability. | who presented with their partners were assigned to the same group. There's a 1/3 chance to be assigned to the pay-it-forward arm (the other two arms were pay-as-you-want and standard of care). If individuals would like to be tested, they would be tested right away on site.<br>The program ran for approximately one month. |
| 10 | Monitoring and evaluation strategy | Through qualitative informant interview.<br>"Polymerase chain reaction (PCR) techniques allowed the researchers to evaluate changes in the bug's food source after housing improvement, thereby confirming a reduced risk of human-vector contact."<br>"Infestation rates decreased dramatically... Spatial analysis<br>of the before and after distribution of vectors." | Data are routinely collected on selected service provision indicators as well as pricing indicators, for better service provision and for sustainability. | This program was carried out as a randomized controlled trial. The process of design, development, implementation and evaluation were carefully monitored and documented. |
| 11 | Setting | The initiative began in four villages and was later scaled up to more than 17 villages in three different countries with diverse ecosystems and ethnic populations | The ITWA diagnostic services are provided in remote and underserved districts in Uganda. Starting from 1 district, growth has continued to at least 6 districts. | This takes place in community-based HIV testing centers in major cities in China (Guangzhou and Beijing). |
| 12 | Adaptability | "The housing improvement strategy and other components of the intervention in the field were then implemented and evaluated. This test provided visibility to the changes that the intervention generated in the homes and in the daily lives of communities, and provided the bases to replicate, implement and scale up the innovation in neighboring countries including El Salvador, Honduras and Nicaragua." | Since its inception, the ITWA program has been expanded both in terms of geographic areas and the services they provide. The program was expanded to six other districts and a total of 11 health facilities by 2016. Wider scale up is envisioned over the next 5 years. Ultrasound sonography was extended to include echocardiography in selected areas. | Pay-it-forward strategy has the potential to be adapted to other context other than the current one. The program was designed with several aspects to enhance generalizability to other community-based testing sites: no doctors were involved in implementation, protocols were streamlined into routine services, and messaging was simplified. Whether the current program can be adapted to more resource-constrained settings need to be further explored. |
| 13 | Financing | Deployed program through international donors. International Development Research Centre (IDRC) of Canada, funded the development of the innovation and supported the scale up to El Salvador and Honduras (2011); the Japanese International Cooperation | Funding is a combination of grants (Phillips, Grand Challenges) as well as minimal client contributions for the service. | The program received funding support from the US National Institutes of Health; the Special Program for Research and Training in Tropical Diseases sponsored by UNICEF, UNDP, World Bank and WHO; the National Key Research and Development Program of China; Doris Duke Charitable Foundation; and the Social Entrepreneurship to Spur |
|  |  | Agency (JICA) funded the transfer of the program to Nicaragua (2014). |  | Health Global. |
| 14 | Social impact | Eco-social model. Three processes emerged, giving shape to this experience and contributing towards interdisciplinarity, intersectorality and community empowerment. These three processes generated a multidisciplinary research team of dynamic partners in governmental, NGO agencies, academia and the community. These processes were not just methodological choices and outcomes of an eco-health approach, but will also be crucial to future social innovations in health. | The social impact includes: improved maternal and health outcomes which directly impact wellbeing of families; increased number of women seeking antenatal care; and increased husband/partner involvement in ANC services. With increased awareness, families and husbands became interested in seeing their unborn child through ultrasonography and preparing for the delivery of the baby. | The program promoted community engagement in health services. In China, men who have sex with men still face social stigmatization and may face difficulties visiting the clinic for sexual health testing services. By partnering with community-based organizations, the program was able to not only provide affordable testing resources, but also empower the community partners to provide more health services to their community. The pay-it-forward action could also build collective agency and social cohesion.<br>From a policy perspective, this type of program could also be useful as a temporary measure to generate testing demand and build trust in new services, before the introduction of more comprehensive public-funded programs. |
| 15 | Health impact | Infestation rates decreased dramatically inside homes and as long as the walls were kept smooth and without crevices, the triatomine bug was unable to establish itself and reproduce within the households. Spatial analysis of | ITWA has expanded to 11 rural health facilities in Uganda and has trained over 150 health workers and conducted over 200,000 ultrasound scans since 2010. Data are used to aid health care decision making for the individual pregnant woman as | Pay-it-forward strategy increased STI testing. 56% men in the pay-it-forward program agreed to receive the gonorrhea and chlamydia test, compared to 46% in the pay-as-you-want group and 18% in the standard-of-care group. |
|  |  | the before and after distribution of vectors [21] substantiated this change. Actual incidence of Chagas was not measured | well as at the specific health facility level. ITWA reports that results of obstetric ultrasound scans have contributed to improved management in about 23% of the total pregnancies. |  |
| 16 | Limitations | First, the period of time for researchers to learn about the initiative and conduct interviews with the communities and other partners was short. Second, the household improvement experience for the control of Chagas disease has been transferred to other countries, but in this case study only the Guatemala initiative was considered - therefore these results may not be generalizable to other contexts. Third, the researchers/authors recognize that evaluation of the cost-benefit relationship of the intervention could contribute to the replicability and sustainability of social innovation in health initiatives. | Not listed | First, the program was examined in two metropolitan cities in China and making inferences to other settings should be done with caution. Second, this program was evaluated in a research context rather than a practice one. The cost-effectiveness analysis used a short-term time zone and did not calculated the disability-adjusted life-years averted or quality-adjusted life-years gained. |
| 17 | Strengths | Using an intersectoral approach, much more than just health outcomes were achieved. | Through task-shifting and development of e-health/telemedicine ultrasound radiology service, the ITWA program made it possible for rural | Compared to the conventional approach, pay-it-forward strategy significantly increased testing uptake and were able to reach more members of key population. The program made |
|  |  |  | pregnant women to receive timely, affordable care closer to home.<br>The business model and implementation strategy focus on self-sufficiency and sustainability, which together are necessary for scaling up this innovation. | gonorrhoea and chlamydia testing more affordable and accessible. |

## Discussion

This research checklist will help to democratize research in social innovation in health and enhance the rigor of research on social innovation in health. It is intended for research on social innovation in diverse global settings, especially LMICs. The research checklist will help to structure research studies and provide guidance for routine monitoring and evaluation related to social innovation in health. Our research checklist extends the literature by focusing on social innovation in health, including iterative feedback from end-users at multiple steps, and using inclusive digital methods that are well adapted for the COVID-19 era.

Our crowdsourcing open call and digital hackathon provided new methods for inclusive end-user feedback, including end-users in LMICs. The process of consensus development is typically driven by experts and some have criticized this process for exclusion of end-users and experts in LMICs. Crowdsourcing open call methods have been used in other health research projects to aggregate wisdom from diverse groups of people.(19) The process involved end-users at all stages of the project, including the modified Delphi process that finalized the checklist. Given the recognized importance of end users in health, our process for consensus development may be relevant to other guideline development at the national or global level.

Our digital hackathon provided an opportunity to transition an in-person method to a series of online workshops. Most hackathons to date have focused on intense in-person collaboration. Potential benefits of the digital hackathon approach include broader inclusion of individuals who would not have been able to join an in-person event, increased time between events to process information and do additional research, and increased capacity to allow real-time participation from people across multiple time zones. We were surprised that despite substantial COVID-19 related competing priorities, the digital hackathon format was effective in identifying consensus.

Our research checklist hackathon process has several limitations. First, the field of social innovation is still emerging and many programs that we would classify as social innovation are not framed this way. Second, the open call required internet access so those without Internet access were not able to participate in the initial open call; alternative methods to solicit ideas and contributions (e.g., unstructured supplementary service data) could increase contributions from people without internet access. Third, we only accepted submissions in English. However, previous global crowdsourcing open calls suggest that when all six official languages of the WHO are options for submissions, greater than 90% are in English.(20)

This research checklist has implications for research and policy. From a research perspective, this checklist will help people in diverse settings to design, implement, and disseminate social innovation in health research. Further research is needed to understand how to measure social innovation in health. Our research checklist raises questions about optimal methods for designing, implementing, and disseminating social innovation in health research. From a policy perspective, our digital hackathon provides an efficient method for collaborative consensus development that is well suited to the COVID-19 era. This could be relevant to policymakers and health leaders organizing consensus processes.

## Conclusion

This 17-item social innovation in health research checklist is the first of its kind and we hope that it will lead to better health and social outcomes through more complete and transparent reporting of the development, implementation, and evaluation of social innovations in health. This research checklist can be used before, during, and after co-creating social innovations in health. Use of the research checklist will help to increase end user and stakeholder engagement, increase the rigor of monitoring and evaluation strategies, consider plans for sustainability, and better determine social and health impacts of social innovation. We hope that researchers, innovators and partners are able to learn more about the processes and results of social innovation in health research projects from each other and that this will drive improved social and health outcomes.

## Supporting information

Social Innovation Research Checklist

## Data Availability

Data sharing is not applicable to this article.

## Acknowledgements

We would like to thank all who contributed to the open call and participated in the Delphi surveys. We also thank Larry Han for his helpful feedback on an earlier version of this manuscript.

## Contributors

EK and EC contributed equally to this manuscript. EK, EC, JT, BH conceptualized the research question. KA, SK, AJO, BH provided feedback on the second version of the manuscript. JF provided guidance on framing social innovation and UA helped define social innovation. PA and IW provide technical advice on social innovation in research. YEK, SP, JT planned and organized the crowdsourcing open call and the Delphi process. All co-authors drafted the manuscript and approved of the final version.

