## Supplementary material for "Social innovation research checklist: A crowdsourcing open call and digital hackathon to develop a checklist for research to advance social innovation in health": Social Innovation Research Checklist

### Social Innovation in Health Research Checklist

| Item | Item No. | Description |
|---|---|---|
| Brief Name | 1 | The title or abstract identified of this social innovation in health research study. |
| Problem | 2 | Describe the current context, background and problem addressed by the social innovation from the perspective of the end user. |
| Rationale | 3 | Describe the rationale for the social innovation, including factors that show a change is needed from the perspective of the end user. |
| Social Innovation | 4 | Describe the key components of the social innovation. This could be accompanied by a detailed description, a photograph, or a figure. Describe each of the processes, activities, and elements used in the social innovation, including any enabling or supporting activities. |
| End Users | 5 | Describe the end users of the social innovation in health. Describe how end users are also direct or indirect beneficiaries of the social innovation. |
| Stakeholder Involvement | 6 | Describe how local stakeholders, including end users, are involved in design, development, implementation, and evaluation of the social innovation in health. In addition, describe the role of marginalized/vulnerable individuals or groups (e.g., people with disability or others as defined by the innovators) in these processes. |
| Inputs | 7 | Describe any physical, digital or informational materials used or distributed during training, delivery and/or implementation of in the social innovation; provide information on where the materials can be accessed† (e.g. online, appendix, URL). |
| Provider | 8 | For each category of the social innovation provider (e.g. community member, trained layperson, other individual), describe their expertise, background, role and any specific training given. |
| Implementation Strategy | 9 | Describe the implementation strategy for the social innovation and whether it is delivered individually, as a group, or partnership. Describe the level of external resources for implementation (e.g., internet access). Describe the frequency and |

| | | |
|---|---|---|
| | | duration of the social innovation delivery. |
| Monitoring & Evaluation Strategy | 10 | Describe what is measured, how, and when as part of monitoring and evaluation. This includes measurement of health, social, and other impacts. |
| Setting | 11 | Describe the population, type(s) of location(s) where the social innovation is delivered, including any necessary social, political, cultural, environmental or other contextual issues. Describe at what level the innovation is implemented (e.g., district, subdistrict, village). This includes a description of the online setting for online social innovation. |
| Adaptability | 12 | Consider how the social innovation could be adapted, scaled up, or used in contexts other than the one described, if appropriate. |
| Financing | 13 | Describe how the social innovation in health has been funded at design, development, implementation, and evaluation stages. Describe how the social innovation could generate revenue (if applicable) or be institutionalized (if applicable) in order to be sustained in the future. |
| Health Impact | 14 | Describe the health impact of the social innovation over a period of time and the methods to assess health impact. Health is defined broadly here according to the WHO definition. |
| Social Impact | 15 | Describe the non-medical impact of the social innovation over a period of time. This could be impact on the environment, social changes, or other non-medical impact (e.g. lessons learned, new processes that emerged from the project, new relationships and networks, application of learned processes to other problems). |
| Limitations | 16 | Describe the limitations and potential unintended consequences of the social innovation in health during the design, development, or implementation. |
| Strengths | 17 | Describe how the social innovation in health improves on conventional practice. |
